# Comparative Effectiveness and Cardiovascular Outcomes of Infliximab reference product and Biosimilars in Patients with Inflammatory Bowel Disease

**DOI:** 10.64898/2026.01.25.26344807

**Authors:** Amirah H. Alnahdi, Naueen Chaudhry, Alaa Alshehri, Qing Liu, Mikael Svensson, Ellen M. Zimmermann, Tianze Jiao

## Abstract

**Background:** Anti–tumor necrosis factor (anti-TNF), particularly infliximab, have transformed inflammatory bowel disease (IBD) management, but their high cost imposes a significant economic burden. Infliximab biosimilars were introduced to reduce the unmet needs. Despite the approval of infliximab biosimilars, real-world evidence of cardiovascular safety and effectiveness of infliximab biosimilars is lacking among patients with IBD. In this trial emulation, we compared the effectiveness and cardiovascular safety between patients who initiated infliximab reference product (IFX-RP) and biosimilars (IFX-BP).

**Methods:** Using the Merative Marketscan Research database (2011–2023), we conducted a retrospective cohort study to emulate the target trial where biologic-naïve adults were randomly assigned to initiate IFX-RP or IFX-BP. Primary outcomes included healthcare resource utilization (HRU), and incidence of major adverse cardiovascular events (MACE) over one year. Propensity score matching was applied to mimic the randomization. Both intention-to-treat and per-protocol effects were estimated.

**Results:** After matching, 850 patients (425 per group) were included. HRU was comparable between IFX-RP and IFX-BP groups across outpatient visits, hospitalizations, surgeries, and emergency visit. During follow-up, MACE events were more frequent in the IFX-BP group (9 vs. 3), with an incidence rate ratio (IRR) of 3.04 (95% CI: 0.82–11.23). Although the difference was not statistically significant, consistent directional trends were observed across analyses. Sensitivity analyses supported primary results.

**Conclusion:** Our study found comparable effectiveness between IFX-RP and IFX-BP in routine clinical care. While cardiovascular events were infrequent, the potential signal suggesting increased MACE risk associated with infliximab biosimilars warrants further investigation. Continued pharmacovigilance is essential to ensure the cardiovascular safety of biosimilars.

**Summary:** Infliximab biosimilars, introduced to reduce the economic burden of anti-TNF therapy in IBD, demonstrated comparable real-world effectiveness to the infliximab reference product in a target trial emulation using Merative MarketScan data, while a potential signal of increased cardiovascular risk underscores the need for ongoing pharmacovigilance and further investigation.

**Key Messages:**

- What is known? Infliximab biosimilars demonstrate comparable efficacy and safety to the reference product in IBD, but cardiovascular outcomes remain underexplored.
- What is new here? This U.S. real-world study emulating a target trial found similar effectiveness between infliximab reference and biosimilar products, with a possible trend toward increased cardiovascular risk in biosimilar users.
- How can this study help patient care? Findings highlight the need for continued pharmacovigilance and cardiovascular monitoring when prescribing infliximab biosimilars to optimize safety in IBD management.

## Background

Advanced therapies have transformed IBD management by targeting specific immune pathways in inflammation, offering effective options for patients who do not respond to conventional therapies^(1,2)^. Infliximab is a chimeric monoclonal antibody that binds to and neutralizes tumor necrosis factor-alpha (TNF-α), a key pro-inflammatory cytokine implicated in the pathogenesis of IBD^(3)^. It’s the first biologic approved for treatment of CD and remains a cornerstone in IBD management^(4)^.

Chronic inflammation in IBD is increasingly recognized as a contributor to elevated cardiovascular risk^(5–7)^. TNF-α, a key inflammatory mediator, has been found at elevated levels in patients with heart failure and myocardial infarction and is considered a predictor of future cardiovascular events^(8–10)^. Furthermore, reductions in TNF-α levels has been linked to favorable cardiovascular outcomes^(11)^. Anti-TNF therapies have demonstrated cardioprotective effects in various immune-mediated inflammatory diseases (IMIDs). In rheumatoid arthritis (RA), a meta-analysis reported a 30% reduced risk of CVD with anti-TNF (relative risk [RR]: 0.70; 95% CI: 0.54–0.90)^(12)^. Similarly, in patients with psoriasis, its use was associated with a 20% lower risk of major adverse cardiovascular events (MACE) compared to those treated with topical agents (hazard ratio [HR]: 0.80; 95% CI: 0.66–0.98)^(13)^.

Among patients with IBD, randomized controlled trials (RCTs) of anti-TNF agents have not demonstrated an increased risk of MACE, with limited trials reporting cardiovascular outcomes such as congestive heart failure (CHF) and coronary artery disease (CAD). This limited evidence may be due to the absence of systematically monitoring for MACE as a primary outcome, small sample sizes, and relatively short follow-up durations ranging from 8 to 54 weeks^(14–19)^. However, observational studies suggest a protective effect on MACE. Two studies used the French national health insurance data found that compared to nonexposed patients, the use of anti-TNF therapy has been associated with a 21% and 25% reduction on acute arterial events (hazard ratio [HR]: 0.79; 95% CI: 0.66–0.95)^(20)^, and recurrent acute arterial events (HR: 0.75; 95% CI: 0.63–0.90)^(21)^, respectively. However, non-exposed patients may receive treatments ranging from 5-aminosalicylic acid, tofacitinib to vedolizumab, which may introduce confounding due to differences in IBD severity.

Despite the clinical benefits, the cost of advanced therapies has emerged as a key concern^(22)^: imposes a considerable burden on patients and the healthcare system, resulting in reduced quality of life, recurrent hospitalizations, and substantial healthcare expenditures-estimated at approximately $8.5 billion in the U.S. in 2018^(23)^. This growing interest in cost-saving alternatives led to the enactment of the Biologics Price Competition and Innovation Act of 2010, which paved the regulatory pathway for biosimilars^(24)^. Under this Act, a biosimilar could be approved with same indication, matched route, dosage form, and strength as the reference biologic if they share similar structure, same mechanism of action, with no clinical meaningful difference^(25–28)^.

Regulatory approval of infliximab biosimilars for IBD was based on extrapolation from clinical trials conducted in other IMIDs. The PLANETAS and PLANETRA trials demonstrated that the infliximab biosimilar CT-P13 had equivalent pharmacokinetics, efficacy, safety, and immunogenicity compared to the reference infliximab in patients with ankylosing spondylitis and rheumatoid arthritis, respectively^(29,30)^. This extrapolation approach initially raised concerns about its applicability to IBD; however, subsequent randomized controlled trials and observational studies have shown no clinically meaningful differences in safety and effectiveness between infliximab reference product (IFX-RP) and infliximab biosimilar products (IFX-BP) though many were limited by relatively small sample sizes^(31–37)^. Nevertheless, the potential impact of these therapies on cardiovascular safety remains an area of active investigation. The NOR-SWITCH trial, a randomized, double-blind, non-inferiority study, found no statistically significant differences in disease worsening or adverse events after switching from IFX-RP to CT-P13 among patients with IMIDs^(31)^. However, cardiovascular outcomes were not stratified by disease type, limiting insights specific to the IBD population^(31)^.

Given TNF-α’s established role in cardiovascular pathophysiology, and the manufacturing complexity of biologics which may introduce potential variations that could impact clinical outcomes of the biosimilars^(38)^. It is crucial to understand the risk profile of these therapies to informing clinical decision-making and optimizing patient outcomes. In parallel, the high cost associated with these therapies continues to raise concerns regarding effectiveness and sequencing healthcare resource utilization (HRU). To address this gap, we emulated a target trial to compare the 1-year cardiovascular safety and effectiveness of IFX-RP versus IFX-BP among biologic-naïve adult patients with IBD.

## Methods

We first specified a hypothetical target RCT comparing the effect of IFX-BP on MACE and health resource utilization with IFX-RP among patients with IBD who were biologic-naive, and then we emulated this target RCT in an observational setting. The protocol of the target RCT and detailed emulation are summarized in Supplement Table 1. The trial emulation can produce results comparable to the target RCT if rigorously implemented^(39–41)^.

### Data source

A retrospective observational study was carried out utilizing administrative claims data from the Merative Marketscan Research database, from January 1, 2011, through December 31, 2023. These databases are de-identified commercial insurance claims that provide detailed records of reimbursed inpatient, outpatient, and prescription services for employees and their dependents who were enrolled in employer-sponsored private health insurance in the United States. The database contains detailed patient-level information, including demographic variables, clinical diagnoses, medical procedures, and prescription drug use. It also documents healthcare utilization and expenditures across inpatient, outpatient, and pharmacy settings. The protocol of this study received scientific and ethical approval from the Institutional Review Board in the University of Florida, USA. Data are available from MarketScan upon request and approval. Analytical codes will be available on GitHub and upon email request.

### Eligibility criteria

Patients entered the base cohort when diagnosed with IBD defined by the presence of the related International Classification of Diseases, 10th Edition (ICD-10) codes in the first inpatient visit or second outpatient visit (complete code lists are provided in the supplementary materials). Eligible patients were aged 18 years or older at base cohort entry date, had continuously enrolled in health insurance, did not experience any MACE, and did not receive any biologics or biosimilars within 1-year before the base cohort entry date. The cohort creation process was shown in Figure 1.

**Figure 1.**
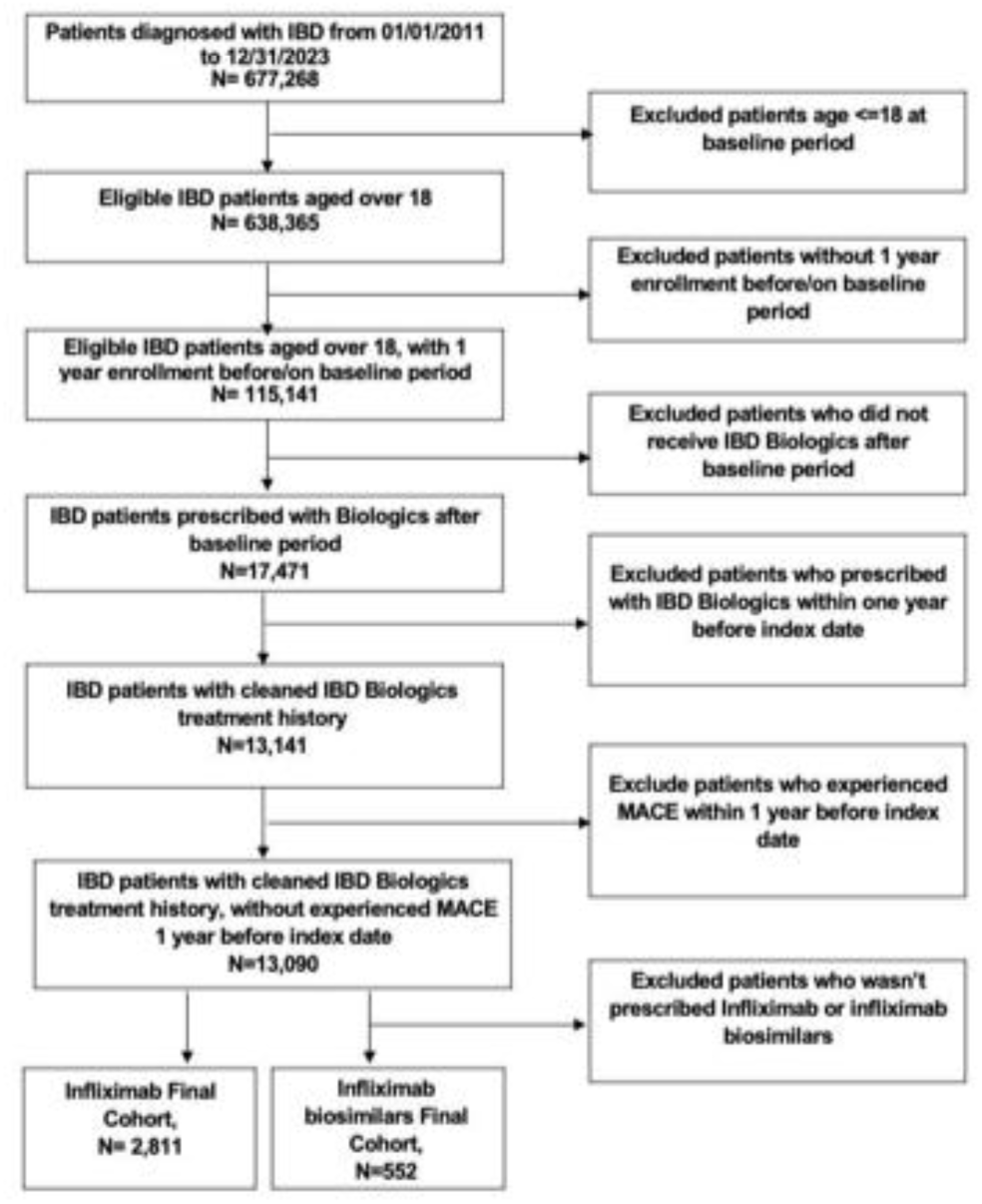
Flow chart

### Treatments

After entering the base cohort, patients joined the study cohort at the date of the first infusion of either IFX-RP or IFX-BP, including infliximab-dyyb, infliximab-abda, and infliximab-axxq, historically names as the index date. Exposure to either treatment was ascertained from two sources: pharmacy claims using National Drug Codes (NDC) or facility-administered medication claims captured via the Healthcare Common Procedure Coding System (HCPCS).

### Covariates

Baseline patient characteristics were assessed during the 12-month baseline period prior to the index date including demographic characteristics (i.e., age, sex), lifestyle factors (i.e., smoking status), comorbidities, IBD medication use, and IBD duration. IBD medication encompassed 5-aminosalicylates, immunomodulators, and steroids, were identified through pharmacy dispensing claims within the baseline period. IBD duration was defined as the time interval between the date of the initial IBD diagnosis and the index date.

### Outcomes

The primary effectiveness outcomes were all-cause and IBD-related HRU defined as utilization of medical services in general or associated with an IBD diagnosis, respectively. As secondary analysis, we further separated the HRU by the type of the location of services received including inpatient hospitalization, emergency room (ER) visits, outpatient visits, and IBD-related surgeries. Surgical procedures included hemicolectomy, colectomy, colostomy, fistulotomy, fistulectomy, ileostomy, ileocecectomy, appendectomy, small bowel resection, and pouch-related procedures. The primary safety outcome was the new onset of MACE following the index date, defined as a hospitalization diagnosed with one of the following conditions: myocardial infarction (MI), stroke, unstable angina, heart failure (HF), or all-cause mortality. Secondary safety outcomes included the individual components of the composite MACE.

### Follow-up

Consistent with the target trial protocol, in the intention-to-treat (ITT) analysis, patients were followed from the date of randomization, index date, until the first occurrence of any of the following events: a study outcome, loss to follow-up (defined by not continuous enrolling in the related health insurance plan), death, or the end of the study period (December 31, 2023). In the per-protocol analysis, patients were additionally censored when discontinuation of assigned treatment or switch to the other studied treatment (IFX-BP or IFX-RP).

### Causal Contrasts of Interest

We assessed the 1-year ITT effect of initiated IFX-BP vs. IFX-RP on the new-onset of MACE, all-cause, or IBD-relate HRU among adult patients with IBD who were biologic-naïve. In the sensitivity analysis, we evaluated the per-protocol effect of initiated and continuously received IFX-BP vs. IFX-RP on the above outcomes.

### Statistical Analysis

To emulate randomization of initiating IFX-RP or IFX-BP, we applied propensity score (PS) matching using the nearest neighbor approach without replacement, with a caliper of 0.25 of the logit of PS. The matching included 30 baseline variables, covering demographic characteristics (i.e., age, sex), lifestyle (i.e., smoking status), cardiovascular risk (indicated by QRISK score), comorbidities, and steroid use (more detail in Table S3). Covariate balance between matched groups was assessed using standardized mean differences (SMDs), with values less than 10% considered indicative of adequate balance.

For the primary effectiveness outcome, mean annual HRU were reported. For the primary safety outcome, the incidence rate ratio (IRR) of MACE was estimated following PS matching. Cox proportional hazards models were used to calculate hazard ratios (HR) with 95% confidence intervals (CI).

In sensitivity analysis, we estimated the per-protocol (PP) effect where we censored patients when they no longer contiguously received the initial biologic therapy. To accommodate imperfect adherence of biologics, we applied a 30-day grace period to identify the continuous use. A p-value of < 0.05 was used to determine statistical significance. All analyses were conducted using SAS Enterprise Guide.

## Results

### Baseline Characteristics

We identified 2,811 and 552 patients initiated IFX-RP or IFX-BP in our cohort before matching (more detail in Figure 1). Some differences were observed between the IFX-RP and IFX-BP groups in several clinical characteristics, including gastrointestinal (GI) bleeding (26% vs. 37%), depression (7% vs. 11%), anxiety (12% vs. 16%), hematological conditions (10% vs. 13%), use of immunomodulators (28% vs. 19%), and 5-aminosalicylic acid (38% vs. 26%). After PS matching, there were 425 patients initiated IFX-RP and IFX-BP, respectively, with comparable demographic and clinical characteristics (more detail in Figure 2). The majority of patients were in the 18-29 and 30-44 age group with ∼51% of males. About 75%, and 74% of patients in the IFX-RP and IFX-BP group received steroids during the 12-month baseline period (more detail in Table 1).

**Figure 2.**
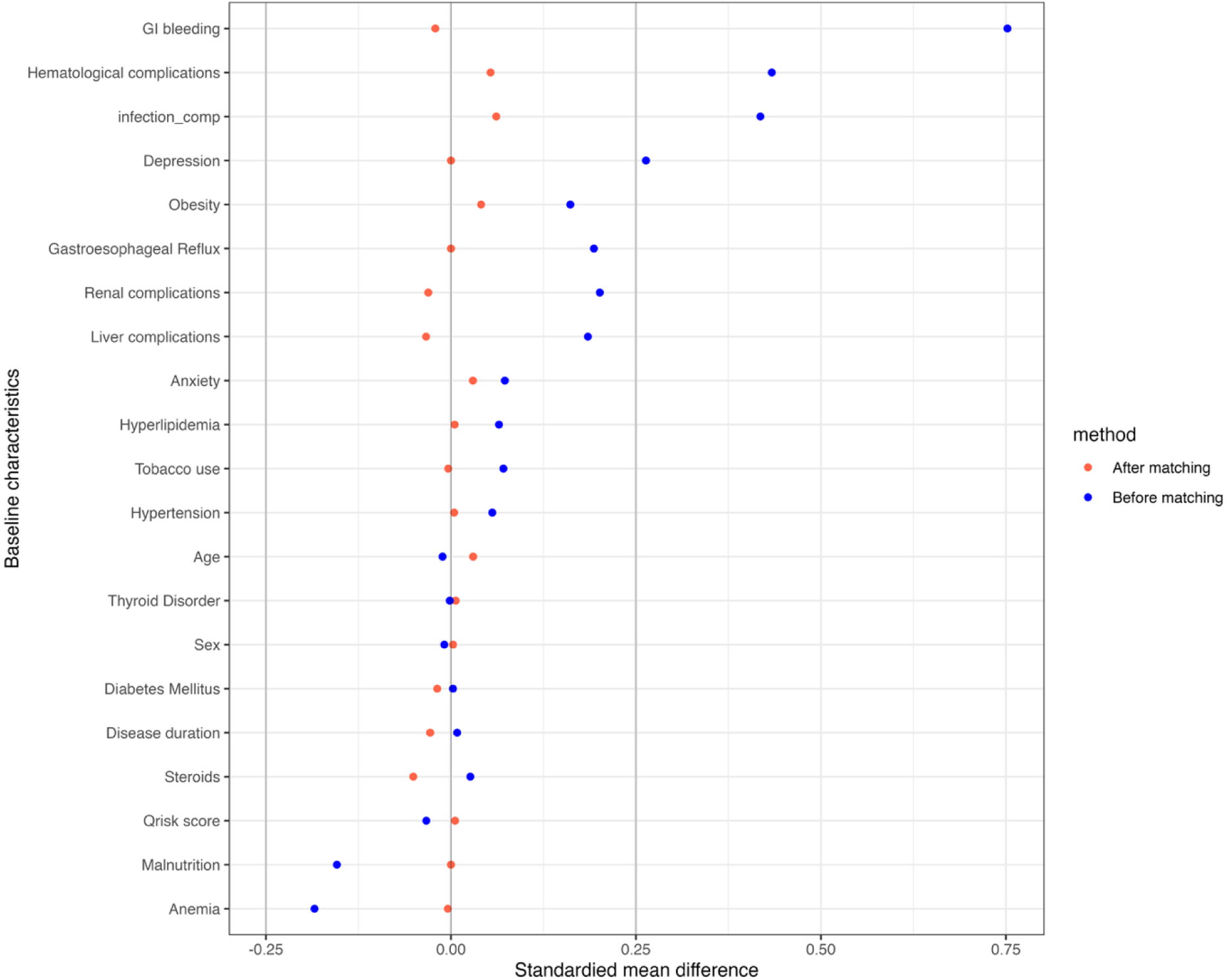
Standardized Mean Differences (SMD) Before and After Propensity Score Matching

**Table 1.**
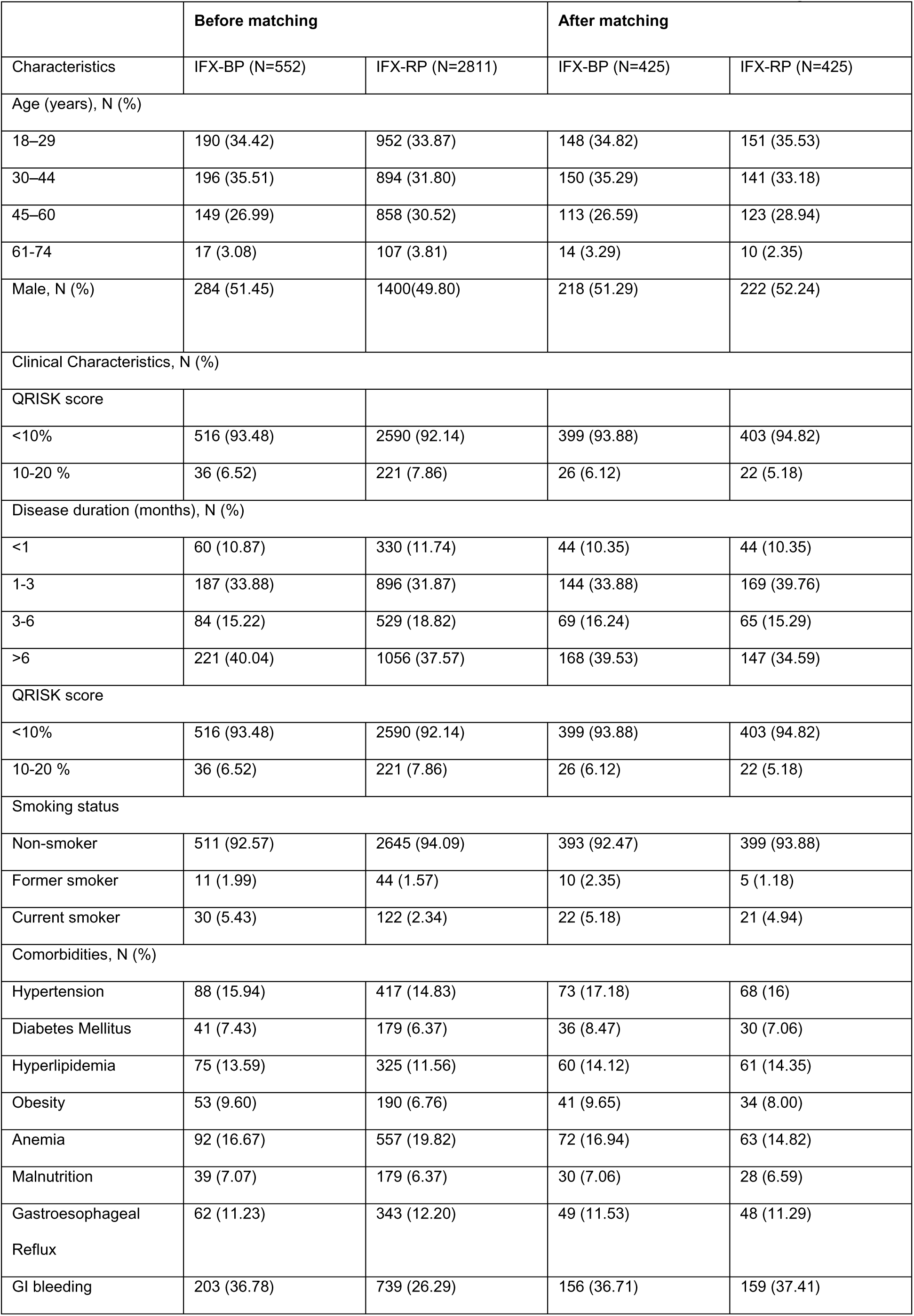
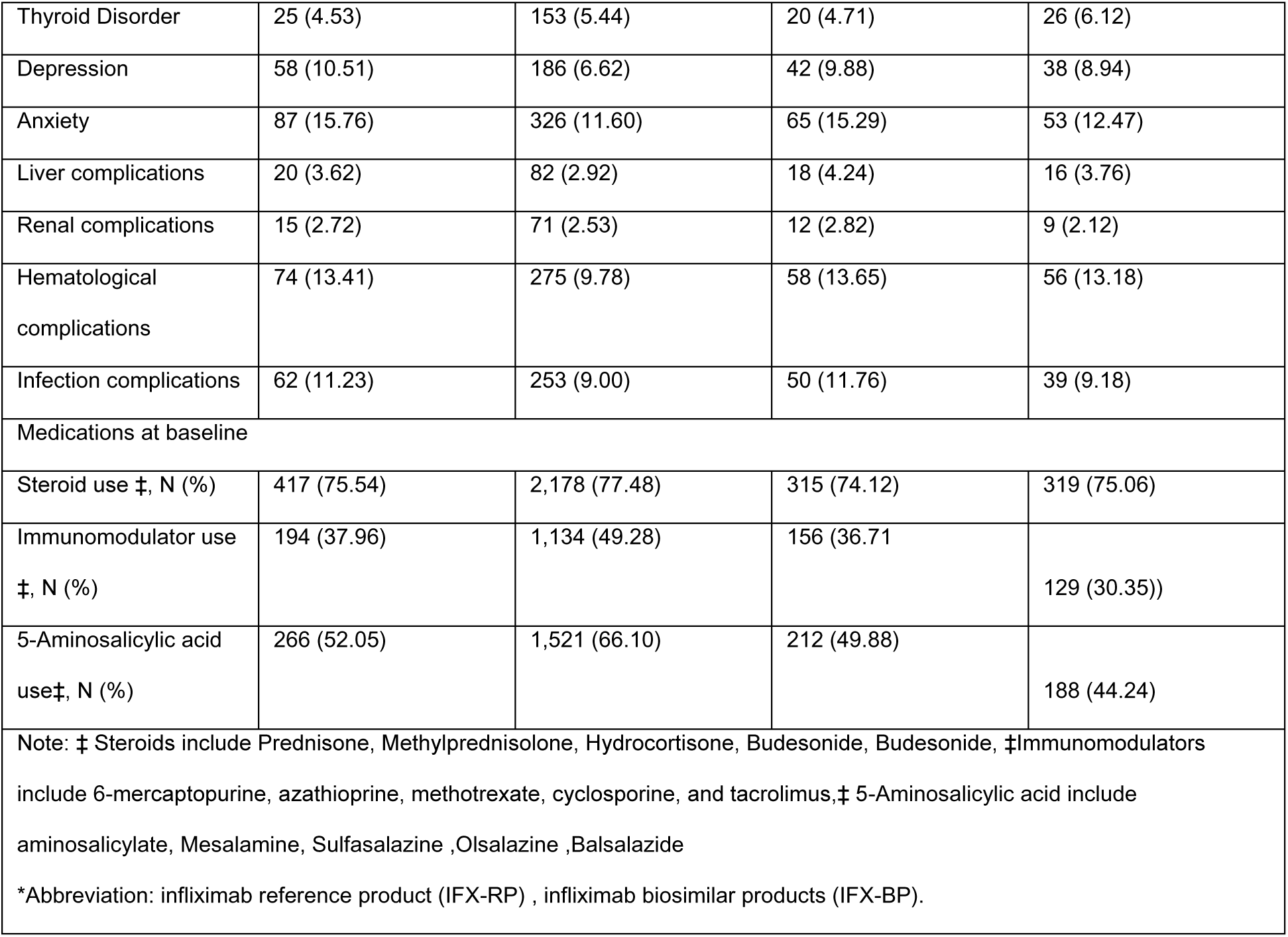
Comparison of baseline characteristics before and after PS matching.

|  | Before matching |  | After matching |  |
| --- | --- | --- | --- | --- |
| Characteristics | IFX-BP (N=552) | IFX-RP (N=2811) | IFX-BP (N=425) | IFX-RP (N=425) |
| Age (years), N (%) |  |  |  |  |
| 18–29 | 190 (34.42) | 952 (33.87) | 148 (34.82) | 151 (35.53) |
| 30–44 | 196 (35.51) | 894 (31.80) | 150 (35.29) | 141 (33.18) |
| 45–60 | 149 (26.99) | 858 (30.52) | 113 (26.59) | 123 (28.94) |
| 61–74 | 17 (3.08) | 107 (3.81) | 14 (3.29) | 10 (2.35) |
| Male, N (%) | 284 (51.45) | 1400(49.80) | 218 (51.29) | 222 (52.24) |
| Clinical Characteristics, N (%) |  |  |  |  |
| QRISK score |  |  |  |  |
| <10% | 516 (93.48) | 2590 (92.14) | 399 (93.88) | 403 (94.82) |
| 10-20 % | 36 (6.52) | 221 (7.86) | 26 (6.12) | 22 (5.18) |
| Disease duration (months), N (%) |  |  |  |  |
| <1 | 60 (10.87) | 330 (11.74) | 44 (10.35) | 44 (10.35) |
| 1-3 | 187 (33.88) | 896 (31.87) | 144 (33.88) | 169 (39.76) |
| 3-6 | 84 (15.22) | 529 (18.82) | 69 (16.24) | 65 (15.29) |
| >6 | 221 (40.04) | 1056 (37.57) | 168 (39.53) | 147 (34.59) |
| QRISK score |  |  |  |  |
| <10% | 516 (93.48) | 2590 (92.14) | 399 (93.88) | 403 (94.82) |
| 10-20 % | 36 (6.52) | 221 (7.86) | 26 (6.12) | 22 (5.18) |
| Smoking status |  |  |  |  |
| Non-smoker | 511 (92.57) | 2645 (94.09) | 393 (92.47) | 399 (93.88) |
| Former smoker | 11 (1.99) | 44 (1.57) | 10 (2.35) | 5 (1.18) |
| Current smoker | 30 (5.43) | 122 (2.34) | 22 (5.18) | 21 (4.94) |
| Comorbidities, N (%) |  |  |  |  |
| Hypertension | 88 (15.94) | 417 (14.83) | 73 (17.18) | 68 (16) |
| Diabetes Mellitus | 41 (7.43) | 179 (6.37) | 36 (8.47) | 30 (7.06) |
| Hyperlipidemia | 75 (13.59) | 325 (11.56) | 60 (14.12) | 61 (14.35) |
| Obesity | 53 (9.60) | 190 (6.76) | 41 (9.65) | 34 (8.00) |
| Anemia | 92 (16.67) | 557 (19.82) | 72 (16.94) | 63 (14.82) |
| Malnutrition | 39 (7.07) | 179 (6.37) | 30 (7.06) | 28 (6.59) |
| Gastroesophageal<br>Reflux | 62 (11.23) | 343 (12.20) | 49 (11.53) | 48 (11.29) |
| GI bleeding | 203 (36.78) | 739 (26.29) | 156 (36.71) | 159 (37.41) |
| Thyroid Disorder | 25 (4.53) | 153 (5.44) | 20 (4.71) | 26 (6.12) |
| Depression | 58 (10.51) | 186 (6.62) | 42 (9.88) | 38 (8.94) |
| Anxiety | 87 (15.76) | 326 (11.60) | 65 (15.29) | 53 (12.47) |
| Liver complications | 20 (3.62) | 82 (2.92) | 18 (4.24) | 16 (3.76) |
| Renal complications | 15 (2.72) | 71 (2.53) | 12 (2.82) | 9 (2.12) |
| Hematological complications | 74 (13.41) | 275 (9.78) | 58 (13.65) | 56 (13.18) |
| Infection complications | 62 (11.23) | 253 (9.00) | 50 (11.76) | 39 (9.18) |
| Medications at baseline |  |  |  |  |
| Steroid use ‡, N (%) | 417 (75.54) | 2,178 (77.48) | 315 (74.12) | 319 (75.06) |
| Immunomodulator use<br>‡, N (%) | 194 (37.96) | 1,134 (49.28) | 156 (36.71) | 129 (30.35)) |
| 5-Aminosalicylic acid<br>use‡, N (%) | 266 (52.05) | 1,521 (66.10) | 212 (49.88) | 188 (44.24) |
| Note: ‡ Steroids include Prednisone, Methylprednisolone, Hydrocortisone, Budesonide, Budesonide, ‡Immunomodulators include 6-mercaptopurine, azathioprine, methotrexate, cyclosporine, and tacrolimus,‡ 5-Aminosalicylic acid include aminosalicylate, Mesalamine, Sulfasalazine ,Olsalazine ,Balsalazide<br>*Abbreviation: infliximab reference product (IFX-RP) , infliximab biosimilar products (IFX-BP). |  |  |  |  |

### Healthcare Resource Utilization (HRU)

Table 2 shows the number of annual HRU for individual patient. On average, patients who initiated IFX-RP experienced 0.21 (SD: 0.93) and 21.03 (SD: 19.00), 0.80 (SD: 1.67) all-cause inpatient, outpatient, or ER visits, respectively. The number decreased to 0.17 (SD: 0.73), 10.88 (SD: 7.18), 0.17 (SD: 0.66) for IBD related inpatient, outpatient or ER visits, respectively. One out of 20 IBD patients who initiated IFX-RP may receive a surgery during 1-year follow-up. No statistically significant differences were observed between patients treated with IFX-RP and IFX-BP across all effectiveness outcomes.

**Table 2.** Annual Healthcare Resource Utilization (HRU), intention-to-treat (ITT) analysis.

| HRU | IFX-BP | IFX-RP | P-value |
| --- | --- | --- | --- |
|  | Mean (SD) | Mean (SD) |  |
| All-cause hospitalization | 0.20 (0.73) | 0.21 (0.93) | 0.87 |
| All-cause outpatient visit | 20.81 (16.51) | 21.03 (19.00) | 0.86 |
| IBD-related hospitalization | 0.17 (0.68) | 0.17 (0.73) | 1 |
| IBD-related outpatient visit | 10.94 (7.40) | 10.88 (7.18) | 0.90 |
| IBD-related surgical procedure | 0.08 (0.50) | 0.05 (0.22) | 0.21 |
| ER visit | 0.71 (1.45) | 0.80 (1.67) | 0.41 |
| IBD-related ER visit | 0.20 (0.68) | 0.17 (0.66) | 0.48 |
| *Abbreviation: Healthcare resource utilization (HRU) , Infliximab reference product (IFX-RP), Infliximab biosimilar products (IFX-BP), Emergency room (ER). |  |  |  |

The per-protocol analysis included patients with a mean follow-up of 6 to 8 months across different outcomes. Follow-up times were comparable between the IFX-RP and IFX-BP groups, with no significant differences observed (p-values ranging from 0.32 to 0.86). The analysis confirmed the findings from the intention-to-treat analysis. (More details in Supplementary Table S4).

### Major Adverse Cardiovascular Events

As shown in Table 3, IFX-BP group experienced more MACE compared to the IFX-RP group (9 vs. 3 events) with elevated incidence rates (2.83 vs. 0.93/100 person-years) during comparable follow-up. The estimated IRR was 3.04 (95% CI: 0.82–11.23). After adjusting for confounders, there was no association between the initiation of IFX-BP and new-onset MACE, though a positive trend was observed (HR: 3.04, 95% CI: 0.82-11.23). Notably, at the end of 1-year follow-up, Kaplan–Meier curve showed lower event-free survival in IFX-BP (97.2%) compared to IFX-RP (99.3%) (log-rank p = 0.082) (Figure 3).

**Figure 3.**
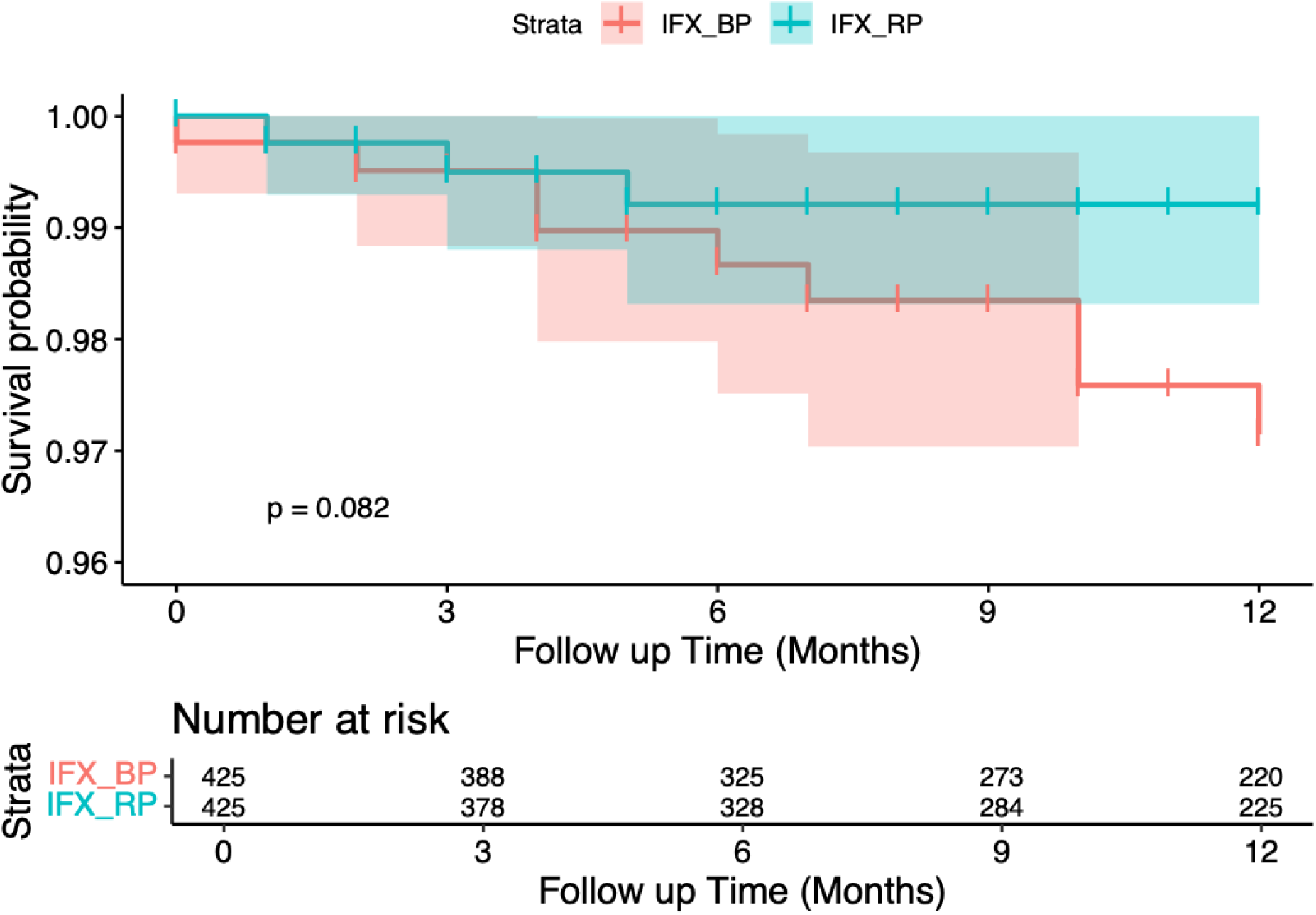
Kaplan-Meier time-to-event analysis within each treatment group at 3-month intervals, intention-to-treat (ITT) analysis

**Table 3.**
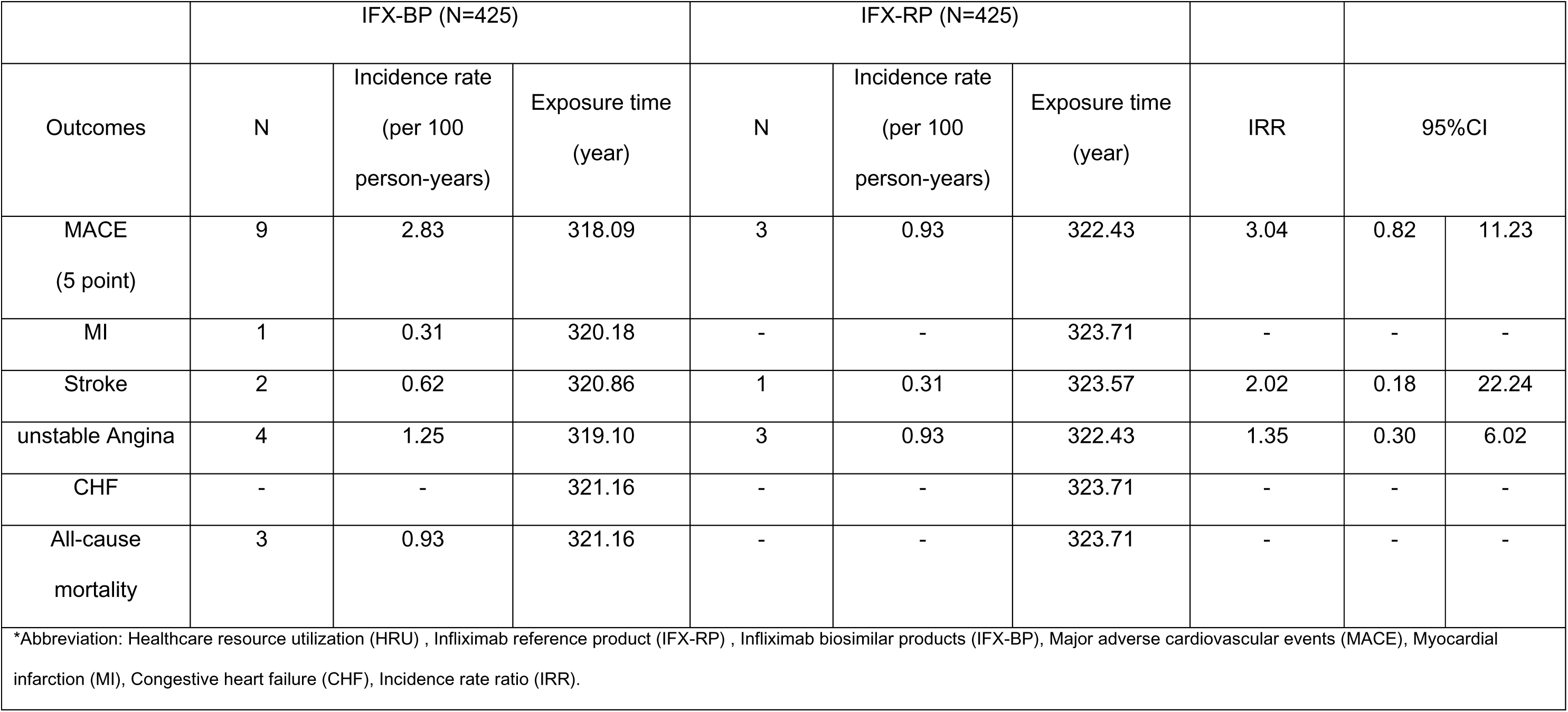
Major Adverse Cardiovascular Events (MACE) incidence rate ratio, intention-to-treat (ITT) analysis.

|  | IFX-BP (N=425) |  |  | IFX-RP (N=425) |  |  |  |  |  |
| --- | --- | --- | --- | --- | --- | --- | --- | --- | --- |
| Outcomes | N | Incidence rate<br>(per 100<br>person-years) | Exposure time<br>(year) | N | Incidence rate<br>(per 100<br>person-years) | Exposure time<br>(year) | IRR | 95%CI |  |
| MACE<br>(5 point) | 9 | 2.83 | 318.09 | 3 | 0.93 | 322.43 | 3.04 | 0.82 | 11.23 |
| MI | 1 | 0.31 | 320.18 | - | - | 323.71 | - | - | - |
| Stroke | 2 | 0.62 | 320.86 | 1 | 0.31 | 323.57 | 2.02 | 0.18 | 22.24 |
| unstable Angina | 4 | 1.25 | 319.10 | 3 | 0.93 | 322.43 | 1.35 | 0.30 | 6.02 |
| CHF | - | - | 321.16 | - | - | 323.71 | - | - | - |
| All-cause<br>mortality | 3 | 0.93 | 321.16 | - | - | 323.71 | - | - | - |

\*Abbreviation: Healthcare resource utilization (HRU) , Infliximab reference product (IFX-RP) , Infliximab biosimilar products (IFX-BP), Major adverse cardiovascular events (MACE), Myocardial infarction (MI), Congestive heart failure (CHF), Incidence rate ratio (IRR).

The event rate of individual components of MACE was low. As shown in Table 3, the IFX-BP group experienced more stroke events compared to the IFX-RP group (2 vs. 1 events) with an HR of 2.07 (95% CI: 0.19–22.79). Similarly, unstable angina events were slightly more frequent in the IFX-BP group (4 vs. 3 events) with an HR of 1.35 (95% CI: 0.30–6.02). Due to low event rates, comparative analyses for MI, CHF, and all-cause mortality were not feasible. Sensitivity analysis to estimate per-protocol effect revealed attenuated differences in the primary outcome (HR: 1.36 (95% CI: 0.30–6.08) relative to the intention-to-treat effect. Notably, the direction of effect for angina was reversed in the per-protocol analysis (HR: 0.68; 95% CI: 0.11–4.09). Low event counts precluded comparative assessment for stroke, MI, CHF, and all-cause mortality (see supplementary materials Table S5).

## Discussion

As the first U.S. real-world study, we successfully emulated the target trial and compared the effectiveness, and cardiovascular safety of IFX-BP versus IFX-RP among biologic-naïve adult patients with IBD. Using the MarketScan database, we found no statistically significant differences in HRU between patients who initiated IFX-RP and those who initiated IFX-BP, after adjusting for baseline characteristics. Meanwhile, we observed an increasing trend on new onset of stroke, unstable angina as well as MACE among those patients who initiated IFX-BP. Yet, none of the association was statistically significant due to the small sample.

This HRU pattern aligns with prior studies showing comparable effectiveness between IFX-BP and IFX-RP initiation in IBD patient^(42–45)^. Moreover, consistent with existing evidence, we observed a shift in IBD-related care from inpatient to outpatient settings, accompanied by reductions in hospitalization and surgery rates over time^(46–48)^. This trend is likely attributable to the adoption of biologic therapies, including infliximab, which have improved the overall management of IBD.

Utilizing a validated machine learning algorithm with 88% positive predictive value (PPV) and 93% sensitivity for mortality detection^(49)^, we observed nine MACE in the IFX-BP group and three in the IFX-RP group, resulting in (HR = 3.04, 95% CI: 0.82–11.23). Individual MACE component analysis revealed consistent patterns despite statistically insignificant due to the sample size. IFX-BP group demonstrated a two-fold increase in stroke risk (HR = 2.07, 95% CI: 0.19–22,79) and a 35% elevation in unstable angina risk (HR = 1.35, 95% CI: 0.30–6.01). Sensitivity analyses that evaluated the per-protocol effect showed consistent directions and magnitudes of effects with some attenuation, which confirmed the robustness of our primary findings. Overall MACE risk remained elevated (HR = 1.36, 95% CI: 0.30–6.08; IRR = 1.35, 95% CI: 0.30–6.02; p = 0.688). Though insignificant, unstable angina risk estimates showed reversal direction (HR = 0.68, 95% CI: 0.11–4.09; IRR=0.67, 95% CI: 0.11–4.03; p = 0.677) than the rest of outcomes. These trends were consistent across hazard, incidence rate ratios in primary and sensitivity analyses, while not statistically significant, suggesting potential cardiovascular safety signals that warrant further investigation.

Indeed, these findings are in line with the NOR-SWITCH trial, which provides the only available cardiovascular evidence to date, reporting one case of myocardial infarction (MI) in the IFX-RP group and one case of each of MI and atrial fibrillation among patients who switched to IFX-BP^(31)^. Chronic inflammation in IBD is increasingly recognized as a contributor to elevated cardiovascular risk^(5–7)^, and reductions in TNF-α levels have been associated with improved cardiovascular outcomes^(11)^. A recent study suggests that, despite equivalent dosing, IFX-BP may not be fully interchangeable with IFX-RP due to pharmacokinetic differences, reporting significantly lower infliximab concentrations in patients receiving IFX-BP, indicating faster drug clearance, potentially linked to lower Fcγ receptor binding affinity^(50)^. This was discussed by Kang et al.^(51)^, who found that IFX-RP exhibited stronger FcγR-IIIa receptor binding affinity and antibody-dependent cellular cytotoxicity (ADCC) than IFX-BP, leading to better efficacy. These findings raise the possibility that minor structural variations, could influence drug behavior and therapeutic effect, potentially explaining the reduced effectiveness and increased trend on cardiovascular outcomes seen with IFX-BP in our findings.

Alternatively, the therapeutic benefits of TNF-α inhibition could be compromised by the development of antidrug antibodies (ADAs), which can lower drug concentrations, diminish efficacy^(52,53)^. While IFX-BP and IFX-RP show similar ADA rates^(31–35)^,individual factors such as genetics, disease type, biologic dose, and patient’s immune response, could influence ADA development^(54)^, potentially explaining the reduced effectiveness seen with IFX-BP in our findings. Future RCTs with long-term follow-up is needed to assess response durability, and long-term safety including cardiovascular outcomes.

There are several strengths of this study. First, in alignment with the principles of target trial emulation, we outlined a clear protocol for the hypothetical trial and detailed the steps used to emulate it using MarketScan database, enhancing the transparency and reproducibility of our study design. Second, we applied propensity score matching rigorously and reported findings with a high degree of transparency. For example, we exactly matched on the time from IBD diagnosis to treatment initiation to reduce time-related bias. Third, we used an incident new-user design to minimize bias from prior drug exposure and more closely approximate the conditions of a randomized controlled trial. Finally, to assess the consistency and robustness of our findings, we conducted a series of secondary (i.e. individual components of the composite MACE) and sensitivity analyses (i.e. per-protocol effect). Majority of these analyses showed consistent results as primary analysis ensuring reliability of our findings.

This study also has several limitations. First, we relied on diagnosis and procedure codes to define covariates and outcomes, which may lead to misclassification. However, these coding algorithms have been widely validated and are routinely used in epidemiologic research. For example, administrative claims–based definitions for 5-point MACE have demonstrated good validity, with reported PPV ranging from 70% to 90% across individual components in various validation studies^(55–58)^. Second, the MarketScan database lacks clinical detail, such as laboratory values (i.e., C-reactive protein and erythrocyte sedimentation rate) and disease activity measures, which may limit our ability to fully adjust for disease severity. Third, medication exposure was defined based on prescription fills rather than actual drug administration, which may lead to overestimation of true exposure to either IFX-BP or IFX-RP and underestimate the rate of MACE. However, we expect majority of patients took the medication due to the medication cost and relief on symptoms, and it should be consistent between the IFX-BP and IFB-RP group. Fourth, as patients were covered by the employer-sponsored insurance, elderly individuals are underrepresented in our cohort, limiting the applicability of our findings to this vulnerable population. Finally, despite rigorous propensity score matching, the possibility of residual confounding cannot be ruled out. Nonetheless, the results of multiple secondary and sensitivity analyses, including estimation of the per-protocol effect, were consistent with the primary findings, supporting the robustness of our conclusion.

## Conclusion

This real-world study found comparable effectiveness between IFX-RP and IFX-BP and provide preliminary evidence suggesting a potential increased cardiovascular risk associated with the use of infliximab biosimilars. While not definitive, these findings highlight a possible signal that hasn’t been adequately explored in previous studies and underscore the need for continuously monitoring the cardiovascular safety of biosimilars.

## Acknowledgements

Amirah H. Alnahdi is supported by the Ministry of Health of Saudi Arabia and the Saudi Arabian Cultural Mission in the USA.

## Data Availability Statement

The data supporting the findings of this study are provided in the supplementary materials.

## Abbreviations

anti-TNF: anti–tumor necrosis factor
IBD: Inflammatory bowel disease
IFX-RP: Infliximab reference product
IFX-BP: infliximab biosimilar product
HRU: Healthcare resource utilization
MACE: Major adverse cardiovascular events
IRR: Incidence rate ratio
CD: Crohn’s disease
UC: Ulcerative colitis
IMIDs: Immune-mediated inflammatory diseases
CVD: Cardiovascular disease
CHF: Congestive heart failure
CAD: Coronary artery disease
MI: Myocardial infarction
HF: Heart failure
GI: Gastrointestinal
TNF-α: Tumor necrosis factor-alpha
ADAs: Antidrug antibodies
ADCC: Antibody-dependent cellular cytotoxicity
RCT: Randomized controlled trial
ITT: Intention-to-treat
PP: Per-protocol
HR: Hazard ratio
CI: Confidence interval
RR: Relative risk
PS: Propensity score
SMD: Standardized mean difference
ER: Emergency room
NDC: National Drug Code
HCPCS: Healthcare Common Procedure Coding System
ICD-10: International Classification of Diseases, 10th Edition
PPV: Positive predictive value
BPCI Act: Biologics Price Competition and Innovation Act
FcγR: Fc gamma receptor

